# Genetics of symptom remission in outpatients with COVID-19

**DOI:** 10.1101/2021.02.24.21252396

**Authors:** Marie-Pierre Dubé, Audrey Lemaçon, Amina Barhdadi, Louis-Philippe Lemieux Perreault, Essaïd Oussaïd, Géraldine Asselin, Sylvie Provost, Maxine Sun, Johanna Sandoval, Marc-André Legault, Ian Mongrain, Anick Dubois, Diane Valois, Emma Dedelis, Jennifer Lousky, Julie Choi, Elisabeth Goulet, Christiane Savard, Lea-Mei Chicoine, Mariève Cossette, Malorie Chabot-Blanchet, Marie-Claude Guertin, Simon de Denus, Nadia Bouabdallaoui, Richard Marchand, Zohar Bassevitch, Anna Nozza, Daniel Gaudet, Philippe L L’Allier, Julie Hussin, Guy Boivin, David Busseuil, Jean-Claude Tardif

**Author notes:** Co-corresponding authors. Marie-Pierre Dubé, Montreal Heart Institute, 5000 Belanger Street, Montreal, H1T 1C8, Canada., Jean-Claude Tardif, Montreal Heart Institute, 5000 Belanger Street, Montreal, H1T 1C8, Canada.

## Abstract

We conducted a genome-wide association study of time to remission of COVID-19 symptoms in 1723 outpatients with at least one risk factor for disease severity from the COLCORONA clinical trial. We found a significant association at 5p13.3 (rs1173773; *P =* 4.94 × 10^−8^) near the natriuretic peptide receptor 3 gene (*NPR3*). By day 15 of the study, 44%, 54% and 59% of participants with 0, 1, or 2 copies of the effect allele respectively, had symptom remission. In 851 participants not treated with colchicine (placebo), there was a significant association at 9q33.1 (rs62575331; *P =* 2.95 × 10^−8^) in interaction with colchicine (*P =* 1.19 × 10^−5^) without impact on risk of hospitalisations, highlighting a possibly shared mechanistic pathway. By day 15 of the study, 46%, 62% and 64% of those with 0, 1, or 2 copies of the effect allele respectively, had symptom remission. The findings need to be replicated and could contribute to the biological understanding of COVID-19 symptom remission.

## INTRODUCTION

The infectious disease syndrome known as coronavirus disease 2019 (COVID-19) is caused by the severe acute respiratory syndrome coronavirus 2 (SARS-CoV-2) and has become pandemic in 2020. Although most COVID-19 cases result in mild symptoms, some patients suffer severe symptoms, including severe pneumonia and multiorgan failure. There is also great variability in the duration of COVID-19 symptoms, with a majority of patients reporting remission of symptoms within 14 to 21 days in the outpatient setting,^1^ while some patients experience prolonged multiorgan dysfunction and complications that last for 12 weeks or longer.^2^ The study of host genetics can bring valuable support toward a better understanding of the mechanisms underlying COVID-19 and can guide the development of preventive and therapeutic measures to mitigate the health burden of this pandemic. Multiple efforts are underway to study the contribution of host genetics to COVID-19, with a focus on risk of severe COVID-19 outcomes and risk of infection with the SARS-CoV-2 virus.^3-7^ In particular, one genetic locus, 3p21.31, has repeatedly been associated with severe respiratory illness and hospitalisation for reasons of COVID-19.^5,7^ However, few studies have focused on the genetics of symptoms duration and remission in outpatients. Persistence of COVID-19 symptoms is associated with a higher risk of complications, including prolonged hospitalization, and poor quality of life thereafter.^8^

Recently, the COLCORONA randomised clinical trial compared the benefit of low-dose colchicine to placebo in 4488 outpatient individuals diagnosed with a COVID-19 infection who were 40 years or older and with at least one high-risk criterion for severe disease.^9^ The composite primary endpoint of death or hospitalisation for reasons of COVID-19 occurred in 4.7% of patients in the colchicine group and 5.8% of those in the placebo group (odds ratio (OR), 0.79; 95% confidence interval (CI), 0.61-1.03; *P =* 0.08). In a prespecified analysis of 4159 participants who received a diagnosis of COVID-19 confirmed by a polymerase chain reaction (PCR) test, the primary endpoint occurred in 4.6% and 6.0% of patients in the colchicine and placebo groups respectively (OR 0.75; 95% CI, 0.57 to 0.99; *P =* 0.04).^9^ Participants were invited to take part in an optional genetic substudy to this randomized clinical trial, which is presented here. We investigated the genetic predictors of time to remission of COVID-19 symptoms with the aim to gain a better understanding of disease progression in a recently diagnosed outpatient population.

## METHODS

### Study population

COLCORONA was a randomised, double-blind, placebo-controlled trial comparing colchicine (0.5 mg twice daily for the first 3 days and then once daily for 27 days thereafter) with placebo in a 1:1 ratio (NCT04322682). Patients were eligible if they were at least 40 years of age, had received a diagnosis of COVID-19, were not hospitalised, and presented at least one of the following high-risk criteria: age of 70 years or more, obesity (body-mass index of 30 kg/m^2^ or more), diabetes, uncontrolled hypertension (systolic blood pressure > 150 mm Hg), known respiratory disease, known heart failure, known coronary disease, fever of at least 38.4°C within the last 48 hours, dyspnea at the time of presentation, bicytopenia, pancytopenia, or the combination of high neutrophil and low lymphocyte counts. Main exclusion criteria included inflammatory bowel disease, chronic diarrhea, estimated glomerular filtration rate (eGFR) less than 30 ml/minute/1.73 m^2^, severe liver disease, current treatment with colchicine, or a history of significant sensitivity to colchicine. Clinical evaluation visits occurred by telephone at 15- and 30-days following randomisation from March 2020 to January 2021.

At the time of consent to the main COLCORONA study, 3315 participants were asked whether they could be recontacted to take part in the optional pharmacogenomic substudy. Those who agreed were called back by the team to obtain a separate consent for the genetic study and to plan for saliva sample collection. Exclusion details are shown in **Supplementary Fig. 1**. There were 2441 participants recruited into the genetic study. Of the 2249 samples genotyped, 20 were excluded due to <98% genotyping completion rate, 3 samples with discordant sex between clinical and genetic data sets were excluded, one contaminated sample was removed, 16 genetically-determined related family members were excluded and we excluded 232 outliers from the cluster based on 1000 Genomes CEU reference samples (Utah Residents with Northern and Western European Ancestry) according to multidimensional scaling. One patient was excluded from the intent to treat population, 3 never received the study drug, and 142 patients who did not have a confirmatory COVID-19 diagnostic test were excluded, leaving 1855 patients for analysis. Written informed consent was obtained from all participants. The study was approved by the Montreal Heart Institute research ethics committee and complies with the Declaration of Helsinki.

### Endpoint Definitions

The day of the end of COVID-19 symptoms was reported by the COLCORONA trial participants during the follow-up telephone visits that occurred on days 15 and 30 of the study. The event date was set as the study day of last known symptoms, alternatively patients were censored at the date of the patients’ last telephone visit. Hospitalized participants were considered as having ongoing symptoms. There were 132 (7%) participants with missing or incomplete information on reported symptom dates, leaving 1723 participants for the analysis of time to symptom remission. The proportion of participants with missing information in the genetic substudy was comparable to the proportion in the COLCORONA trial (359/4488; 8%). Further investigations for the 132 excluded participants found related research nurses’ notes for 34 patients, 14 had reported symptom initiation after the baseline visit, and 20 had reported having no symptoms. Information on the dates of symptom occurrence was collected, but the specific symptoms themselves were not collected. No deaths were reported in participants to the genetic study, consequently the severity outcome was limited to hospitalisation for reasons of COVID-19 in the 30 days following randomisation.

### Genotyping and imputation

DNA was obtained using the Oragene OG-500 self-collection saliva kits delivered to and collected from the participants’ home while respecting distancing and quarantine restrictions. DNA was extracted from 1 ml of saliva and genome-wide genotyping was performed using 200 ng of genomic DNA at the Beaulieu-Saucier Pharmacogenomics Centre (Montreal, Canada). The Illumina Infinium Global Screening Array (GSA) v3-MD (Illumina, San Diego, CA) including 700,625 genomic markers was used and processed according to the manufacturer’s specifications. BeadChips were scanned using the Illumina iScan Reader and analysed with the data manifest MHI_GSAMD-24v3-0-EA_20034606_C2.bpm, with minor manual cluster adjustment for ADME genes. Plink files were produced by the iaap-cli tool (version 1.1.0-80d7e5b). Intensities, B allele frequency, and log R ratio were extracted using the gtc_convert tool (version 0.1.2). pyGenClean ^10^ version 1.8.3 and PLINK^11^ version 1.07 or 1.9 were used to conduct quality control and genetic data clean-up processes. The genotyping experiment consisted of 28 plates of DNA samples with one Coriell control per hybridization experiment. The pairwise concordance of control samples ranged from 0.99998 to 0.999998 and concordance of their genotypes with expectation from the 1000 Genomes genotype data ranged from 0.9986 to 0.9989.

The completion rate threshold for genotypes and samples was set to 98%. SNPs with genotyping plate bias (n = 213) based on the 96 well plates used to dilute DNA samples were flagged but not removed as the effect of genetic ancestry could not be excluded. Pairwise identity-by-state (IBS) was used to conduct close familial relationship checks. We removed all but one member of related samples based on a selection of uncorrelated SNPs (r^2^ < 0.1). The pairwise IBS matrix was used as a distance metric to identify cryptic relatedness among samples and to identify cluster outliers by multidimensional scaling (MDS). The coordinates of the first two MDS components of each subject were plotted including the genotypes of HapMap CEU, JPT-CHB, and YRI data (unrelated individuals). Outliers from the main cluster overlapping the CEU reference samples (Utah residents with Northern and Western European ancestry from the CEPH collection) were removed according to k-nearest neighbour with a threshold of 1.9σ in pyGenClean (v1.8.3). Principal components were generated based on the final sample selection and used to control for confounding by ancestry in genetic association analyses.^12^

Genome-wide imputation was performed on the TOPMed Imputation Server (version 1.5.7)^13^ using Eagle (version 2.4)^14^ for phasing and Minimac4 (version 1.0.2)^13^ for imputation. Following quality control, 513,036 genetic variants were used for imputation. The imputation provided 308,070,060 (513,036 genotyped and 307,557,024 imputed) genetic from which we retained 43,062,846 (513,012 genotyped and 42,549,834 imputed) variants with a quality value (r^2^) ≥ 0.6, and of which 6,490,603 had a minor allele frequency (MAF) ≥ 5%. Genotype “hard calls” (non-probabilistic best-guess genotype assignations) were generated for individual genotypes with genotype probability scores ≥ 0.80 and otherwise set to missing. The hard calls were used only in analyses stratified by genotype. Chromosomal positions are according to GRCh38.

### Statistical Analyses

Genome-wide association analysis (GWAS) with common genetic variants (MAF ≥ 5%) were conducted using the program genetest version 0.5.0.^15^ Cox proportional hazards regression was used for the GWAS of time from randomisation to end of COVID-19 symptoms (time to remission) in 1723 study participants. Logistic regression was used for the analysis of the severity endpoint of hospitalisation for COVID-19 in 1855 participants. To avoid bias due to population structure, analyses were limited to unrelated individuals of genetically determined European ancestry, which was the largest population group in the sample. Genetic variants were coded according to imputation dosage in (0 - 2) and tested using a 1-degree of freedom Wald test adjusted for age, sex, and the first 10 principal components to control for genetic ancestry, with the addition of the study treatment arm when both arms were included in the analyses. GWAS were conducted using patients from both study arms, as well as stratified by treatment arms. Exploratory sex-stratified analyses are reported in **Supplementary Material**. The GWAS for hospitalisation due to COVID-19 was only performed using both study arms due to the limited number of events. Each GWAS was conducted at the 5 × 10^−8^ significance level to adjust for the multiple testing of genetic variants within each phenotype. No additional adjustments were made to account for the multiple phenotypes or different statistical methods. Results are reported with point estimates and 95% confidence intervals (CI) which are not adjusted for multiple comparisons. The top findings from the GWAS were reproduced using SAS version 9.4. The proportionality of hazards assumption was verified for variants identified in a Cox proportional hazards regression GWAS, and the convergence criterion was met for each considered model. Cumulative incidence plots were obtained using a parametric model fitted with the R package. In order to quantify the effect of the genetic variants upon the duration of symptoms, we fitted a Weibull accelerated failure time model adjusted for age, sex and 10 principal components using the lifereg procedure in SAS 9.4.

### Functional annotation

We first defined credible candidate variants as those located within 500 kb of the leading variants and with *P* values within two orders of magnitude of the lead variant. We used the software GCTA-CoLo^16^ to conduct a conditional analysis to identify independent signals. We used PAINTOR^17^ to identify credible sets of causal variants based on the magnitude and direction of association and the pairwise linkage disequilibrium structure at the loci, and we used RegulomeDB^18^ and DSNetwork^19^ to assign a relative ranking to variants. We used *in silico* functional annotations from the public databases Open Target Genetics^20^ and PhenoScanner^21,22^ to identify potential functional mechanisms and target genes. We tested the colocalization between the COLCORONA GWAS signals and clinically relevant phenotypes using the COLOC R package v3.2-1.^23^ Network analysis of selected candidate genes was conducted using GeneMANIA.^24^ The complete approach is detailed in **Supplementary Material**.

### Data availability

The anonymized patient level data from the COLCORONA trial will be shared via the Vivli (vivli.org) data repository. The patient level genetic data underlying this article cannot be shared to preserve the privacy of study participants. Summary statistics from the GWAS results are available for download and visualisation via PheWeb^25^ at statgen.org/pheweb/colcorona.

## RESULTS

There were 1855 participants available for the genetic study of COLCORONA (**Supplementary Fig. 1**). The baseline characteristics of the participants to the genetic study are shown in **Table 1**. The mean age of participants was 54.1 years, 56.2% were female, mean BMI was 30.2 kg/m^2^, 16.3% had a medical history of diabetes, 32.1% of hypertension, and 29.4% of respiratory disease. There were 1254 (72.8%) participants who reported being free of COVID-19 symptoms during the study follow-up period, and the mean number of days between randomisation and end of COVID-19 symptoms in those participants was 11.7 days. Mean number of days between first symptoms and randomisation was 5.4 days (**Table 1**). Overall, women reported longer symptom duration than men (**Supplementary Table 1**). The COLCORONA primary endpoint of death or hospitalisation for COVID-19 differed significantly between the full trial population and the genetic substudy. Whereas there were 14 (0.3%) reported deaths in the COLCORONA trial, none of the participants to the genetic study died (*P <* 0.001), and there were 229 (5.1%) hospitalisation for COVID-19 in the trial compared to 58 (3.1%) in the genetic study (*P <* 0.001).

**Table 1.**
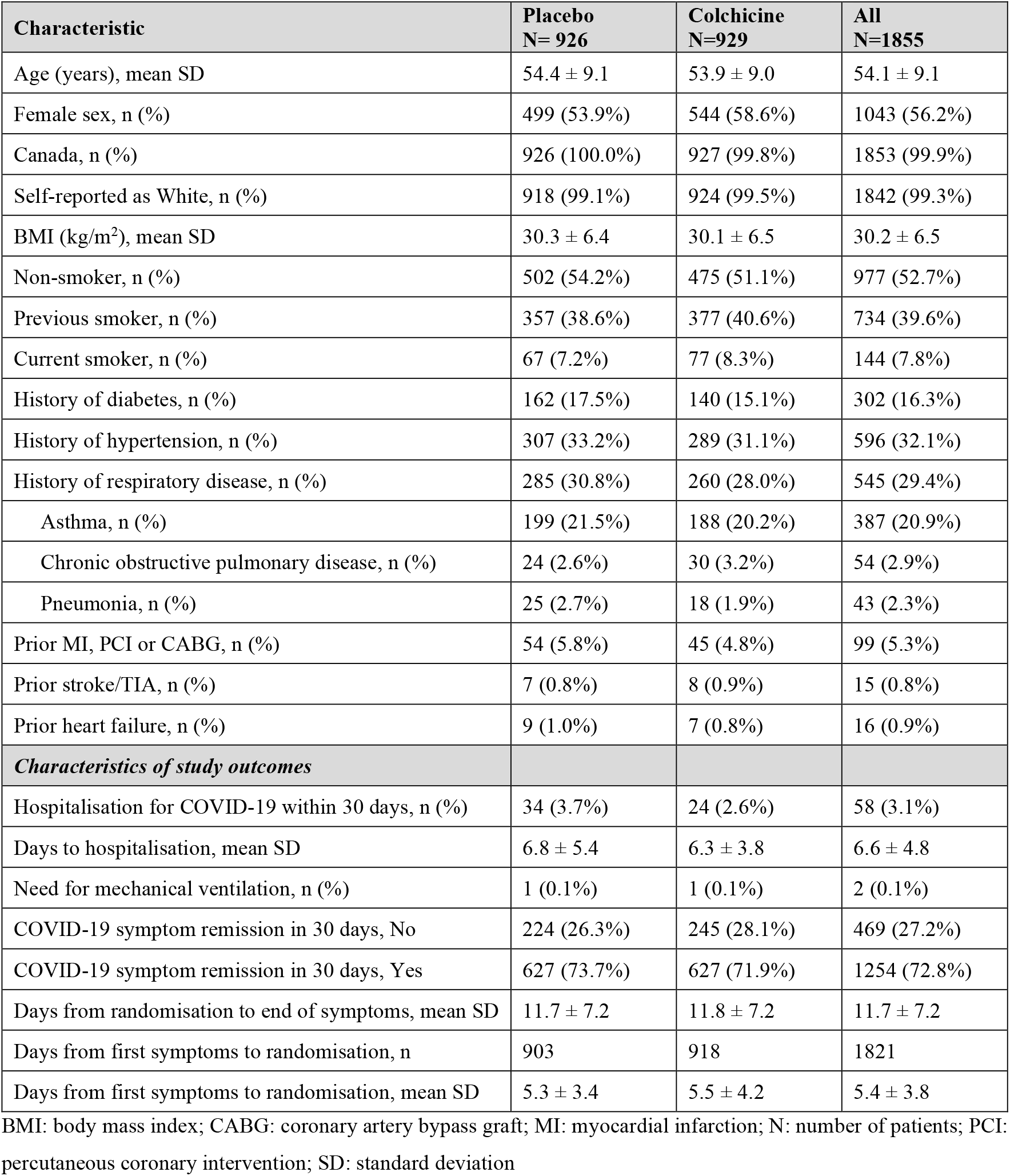
Characteristics of the study population.

| Characteristic | Placebo<br>N= 926 | Colchicine<br>N=929 | All<br>N=1855 |
| --- | --- | --- | --- |
| Age (years), mean SD | 54.4 ± 9.1 | 53.9 ± 9.0 | 54.1 ± 9.1 |
| Female sex, n (%) | 499 (53.9%) | 544 (58.6%) | 1043 (56.2%) |
| Canada, n (%) | 926 (100.0%) | 927 (99.8%) | 1853 (99.9%) |
| Self-reported as White, n (%) | 918 (99.1%) | 924 (99.5%) | 1842 (99.3%) |
| BMI (kg/m<sup>2</sup>), mean SD | 30.3 ± 6.4 | 30.1 ± 6.5 | 30.2 ± 6.5 |
| Non-smoker, n (%) | 502 (54.2%) | 475 (51.1%) | 977 (52.7%) |
| Previous smoker, n (%) | 357 (38.6%) | 377 (40.6%) | 734 (39.6%) |
| Current smoker, n (%) | 67 (7.2%) | 77 (8.3%) | 144 (7.8%) |
| History of diabetes, n (%) | 162 (17.5%) | 140 (15.1%) | 302 (16.3%) |
| History of hypertension, n (%) | 307 (33.2%) | 289 (31.1%) | 596 (32.1%) |
| History of respiratory disease, n (%) | 285 (30.8%) | 260 (28.0%) | 545 (29.4%) |
| Asthma, n (%) | 199 (21.5%) | 188 (20.2%) | 387 (20.9%) |
| Chronic obstructive pulmonary disease, n (%) | 24 (2.6%) | 30 (3.2%) | 54 (2.9%) |
| Pneumonia, n (%) | 25 (2.7%) | 18 (1.9%) | 43 (2.3%) |
| Prior MI, PCI or CABG, n (%) | 54 (5.8%) | 45 (4.8%) | 99 (5.3%) |
| Prior stroke/TIA, n (%) | 7 (0.8%) | 8 (0.9%) | 15 (0.8%) |
| Prior heart failure, n (%) | 9 (1.0%) | 7 (0.8%) | 16 (0.9%) |
| **Characteristics of study outcomes** |  |  |  |
| Hospitalisation for COVID-19 within 30 days, n (%) | 34 (3.7%) | 24 (2.6%) | 58 (3.1%) |
| Days to hospitalisation, mean SD | 6.8 ± 5.4 | 6.3 ± 3.8 | 6.6 ± 4.8 |
| Need for mechanical ventilation, n (%) | 1 (0.1%) | 1 (0.1%) | 2 (0.1%) |
| COVID-19 symptom remission in 30 days, No | 224 (26.3%) | 245 (28.1%) | 469 (27.2%) |
| COVID-19 symptom remission in 30 days, Yes | 627 (73.7%) | 627 (71.9%) | 1254 (72.8%) |
| Days from randomisation to end of symptoms, mean SD | 11.7 ± 7.2 | 11.8 ± 7.2 | 11.7 ± 7.2 |
| Days from first symptoms to randomisation, n | 903 | 918 | 1821 |
| Days from first symptoms to randomisation, mean SD | 5.3 ± 3.4 | 5.5 ± 4.2 | 5.4 ± 3.8 |

BMI: body mass index; CABG: coronary artery bypass graft; MI: myocardial infarction; N: number of patients; PCI: percutaneous coronary intervention; SD: standard deviation

### GWAS for time to remission of COVID-19 symptoms

There were 1723 participants included in the GWAS for time to remission of COVID-19 symptoms. Of those, 1254 (72.8%) reported end of symptoms during the study period and 469 were censored at the date of their last telephone visit. We found two candidate genomic regions significantly associated with time to symptom remission located on chromosomes 5 and 9 (**Fig. 1**). Sex-stratified GWAS did not find additional associations (**Supplementary Material**). We found one significant association signal in the GWAS for time to COVID-19 symptom remission using both study arms at the 5p13.3 locus with the leading variant rs1173773 (*P =* 4.94 × 10^−8^; MAF 0.34). When conditioning on rs1173773, no additional genetic variants remained significant at *P <*5 × 10^−8^ in the region and rs1173773 had the highest probability of being causal according to statistical and functional prioritization. The minor allele (C) was associated with a higher remission rate (hazard ratio (HR) = 1.25, 95% CI 1.15-1.35) as compared to the T allele, irrespective of treatment arm (interaction *P =* 0.18) (**Table 2**). According to survival modeling, 44%, 54% and 59% of patients with the TT, CT and CC genotypes respectively were predicted to have had symptom remission by day 15 of the study (**Fig. 2, Table 3**). Using an accelerated failure time model, we calculated that the C allele contributes to symptom remission with an acceleration factor of 0.84 (95% CI 0.80-0.89) compared to the T allele (*P* = 5.23 × 10^−9^). Compared to patients with the rs1173773-TT genotype, patients with the CC genotype had symptom remission with an acceleration factor of 0.73 (95% CI 0.64-0.82; *P* = 3.89 × 10^−7^), and the CT genotype had an acceleration factor of 0.82 (95% CI 0.75-0.89; *P* = 6.30 × 10^−6^). The rs1173773 variant is located in intron 3 of the most abundant transcript of the natriuretic peptide receptor 3 gene (*NPR3*). However, functional analysis found no evidence supporting a regulatory role for this variant. The rs1173773 C allele that is associated with shorter duration of COVID-19 symptoms was previously found to be associated with greater standing height,^26^ and variants in *NPR3* have also been associated with forced expiratory volume in 1 second (FEV1),^26^ and systolic blood pressure.^27^

**Table 2.**
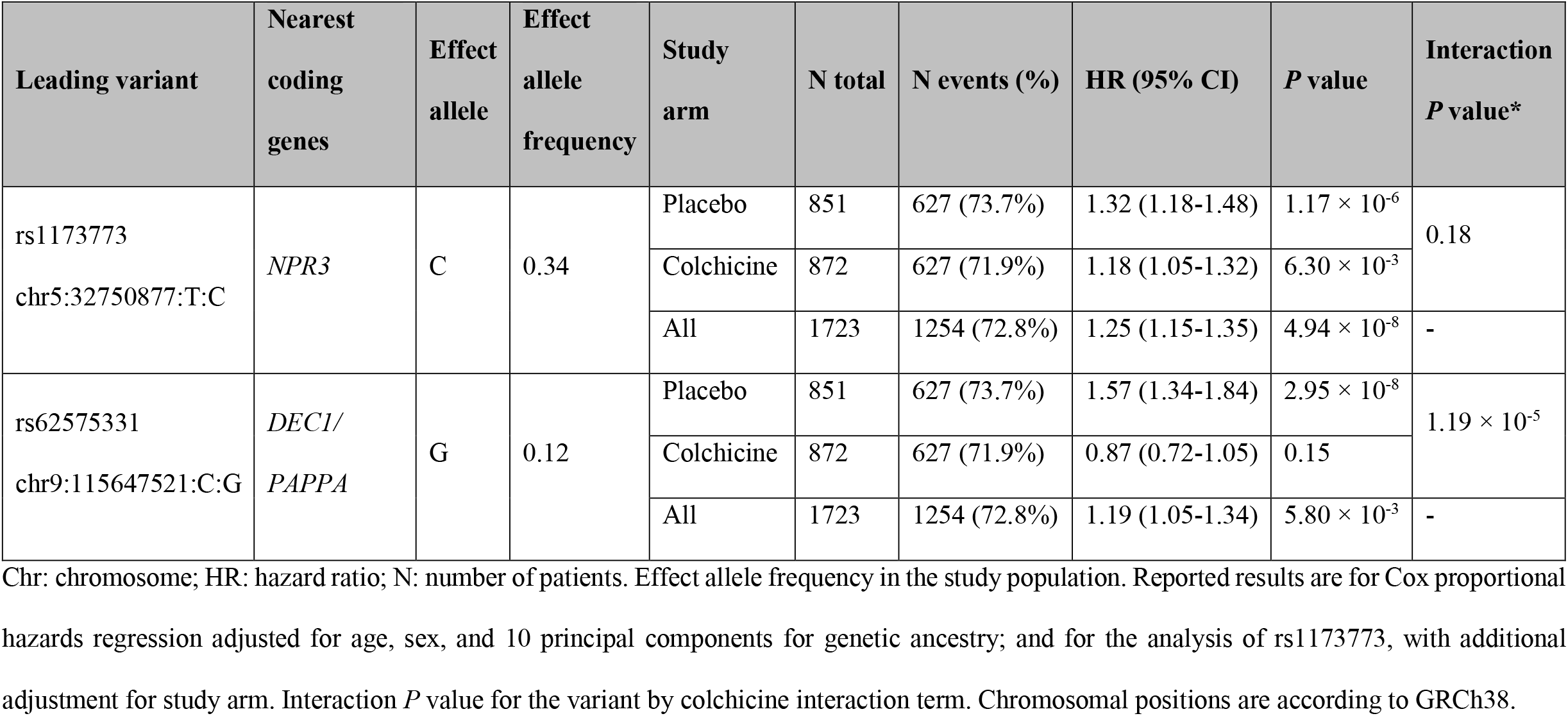
Genetic association results of the leading genetic variants identified in the GWAS for time to COVID-19 symptom remission.

**Table 3.** Estimated cumulative incidence of COVID-19 symptom remission events by genotype groups.

|  | rs1173773 (chr5:32750877:T:C) |  |  |  |  |  |  |  |  |  |  |  |
| --- | --- | --- | --- | --- | --- | --- | --- | --- | --- | --- | --- | --- |
|  | All genotyped |  |  | TT |  |  | CT |  |  | CC |  |  |
| Day | At risk | Remission events | Remission Proportion | At risk | Remission events | Remission Proportion | At risk | Remission events | Remission Proportion | At risk | Remission events | Remission Proportion |
| 0 | 1719 | - | - | 770 | - | - | 734 | 0 | 0.00 | 215 | - | - |
| 5 | 1600 | 173 | 0.10 | 725 | 65 | 0.08 | 680 | 82 | 0.11 | 195 | 26 | 0.12 |
| 10 | 1238 | 404 | 0.33 | 579 | 167 | 0.30 | 513 | 184 | 0.35 | 146 | 53 | 0.36 |
| 15 | 860 | 303 | 0.50 | 426 | 113 | 0.44 | 341 | 139 | 0.54 | 93 | 51 | 0.59 |
| 20 | 712 | 127 | 0.58 | 365 | 59 | 0.52 | 277 | 52 | 0.61 | 70 | 16 | 0.67 |
| 25 | 559 | 162 | 0.67 | 293 | 76 | 0.62 | 217 | 62 | 0.70 | 49 | 24 | 0.78 |
| 30 | 130 | 83 | 0.73 | 71 | 44 | 0.68 | 50 | 31 | 0.75 | 9 | 8 | 0.82 |
|  | rs62575331 (chr9:115647521:C:G) |  |  |  |  |  |  |  |  |  |  |  |
|  | All genotyped |  |  | CC |  |  | CG |  |  | GG |  |  |
| Day | At risk | Remission events | Remission Proportion | At risk | Remission events | Remission Proportion | At risk | Remission events | Remission Proportion | At risk | Remission events | Remission Proportion |
| 0 | 833 | - | - | 664 | - | - | 153 | 0 | 0.00 | 16 | - | - |
| 5 | 772 | 92 | 0.11 | 626 | 62 | 0.09 | 131 | 27 | 0.17 | 15 | 3 | 0.18 |
| 10 | 590 | 195 | 0.34 | 490 | 147 | 0.31 | 91 | 44 | 0.45 | 9 | 4 | 0.42 |
| 15 | 422 | 135 | 0.50 | 358 | 105 | 0.46 | 60 | 26 | 0.62 | 4 | 4 | 0.64 |
| 20 | 345 | 65 | 0.58 | 297 | 55 | 0.55 | 46 | 8 | 0.67 | 2 | 2 | 0.80 |
| 25 | 266 | 87 | 0.68 | 233 | 71 | 0.66 | 31 | 15 | 0.78 | 2 | 1 | 0.88 |
| 30 | 67 | 37 | 0.73 | 56 | 31 | 0.70 | 11 | 5 | 0.81 | - | - | - |

Cumulative incidence estimates were derived from Cox proportional hazards using genotype hard calls with adjustment for age, sex, 10 principal components, and for the analysis of rs1173773, with additional adjustment for study arm. Remission proportion was calculated as 1- symptom persistence probability. Chr: chromosome.

**Figure 1.**
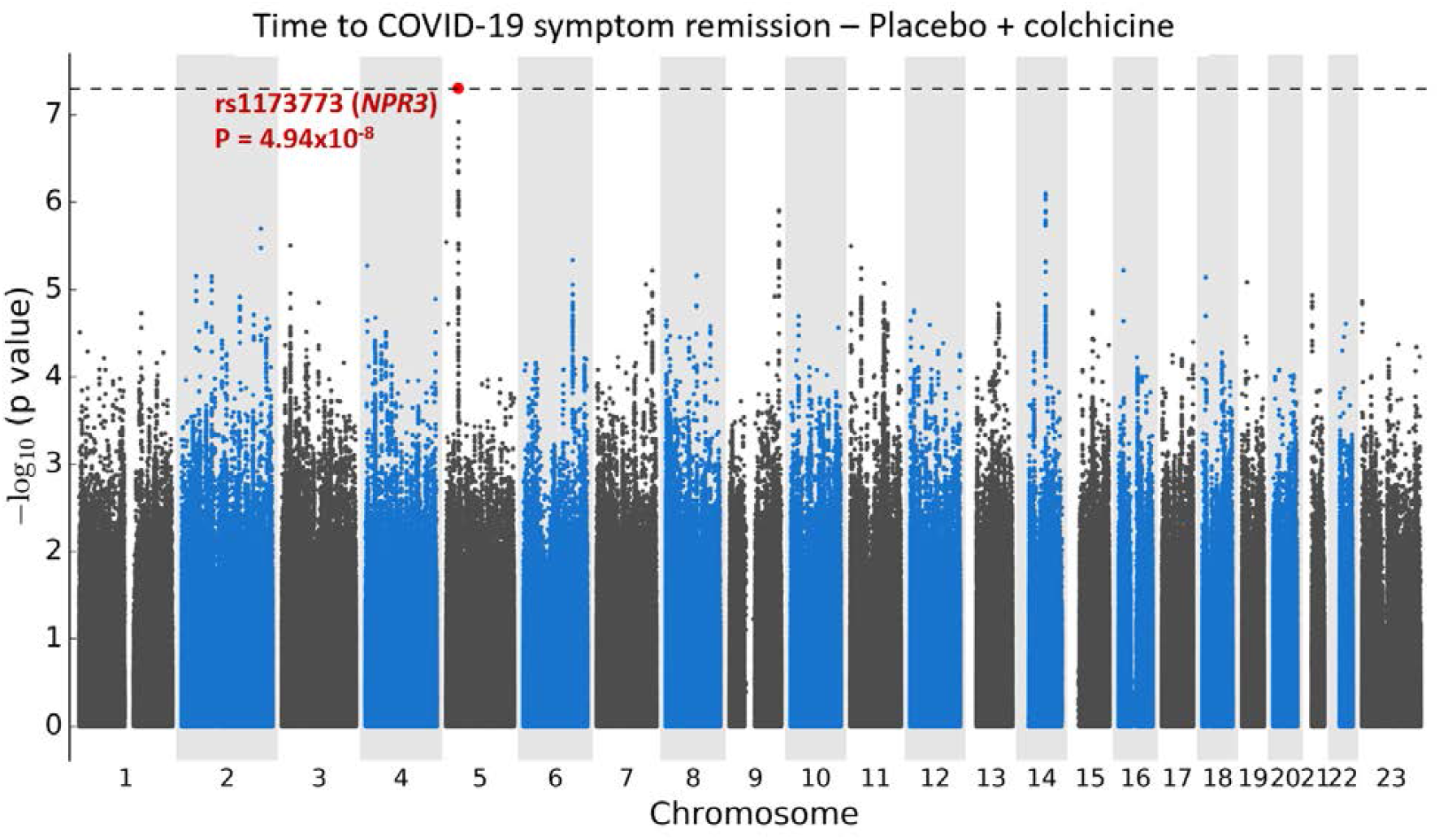
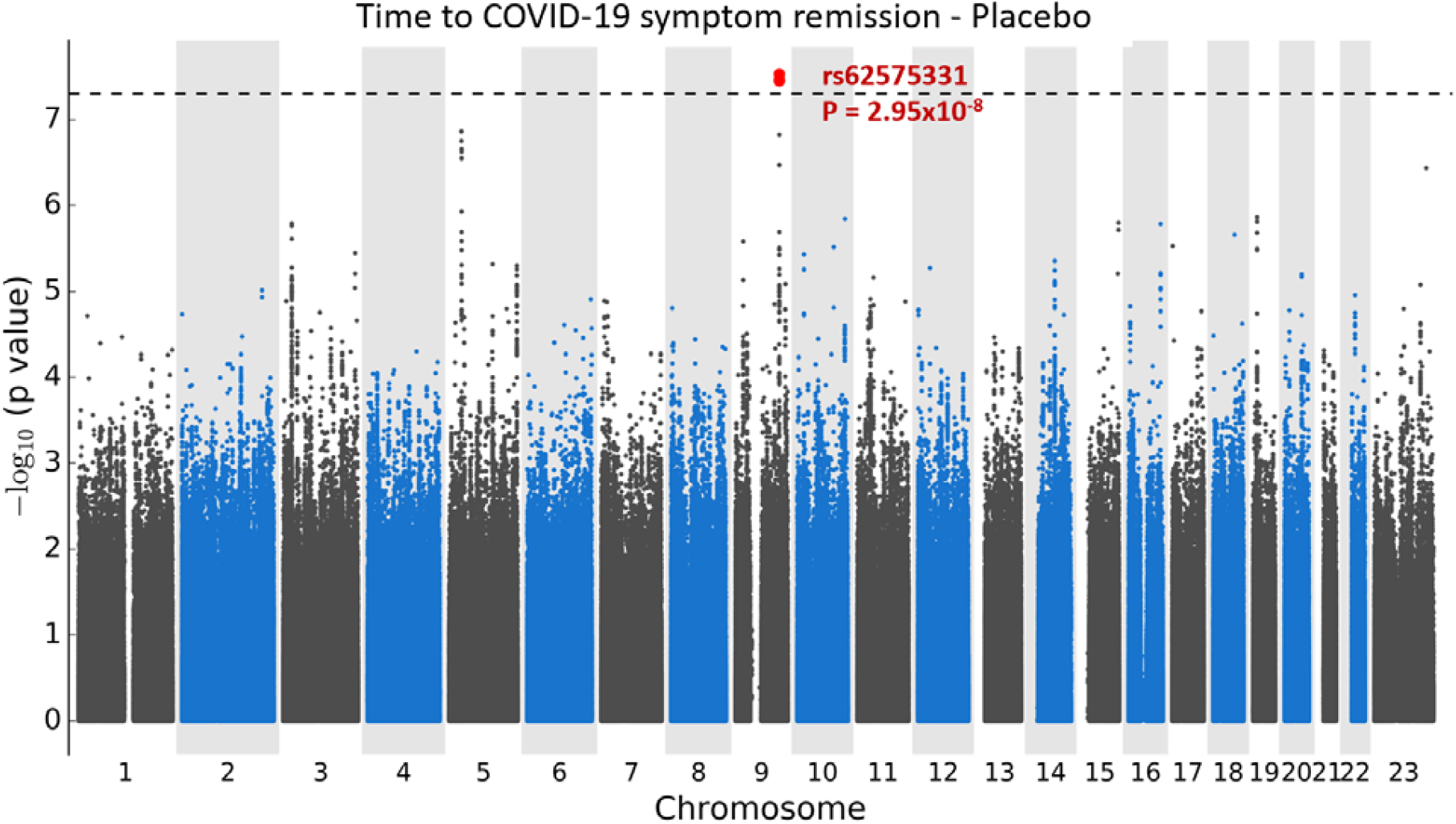
Manhattan plots for the GWAS of time to remission of COVID-19 symptoms. **a**. Using a Cox proportional hazards regression with 1723 subjects from the colchicine and placebo arms of the COLCORONA study, controlling for study arm, sex, age, and 10 principal components for genetic ancestry, with 6,392,715 genetic variants of minor allele frequency ≥ 5%. **b**. Using a Cox proportional hazards regression with 851 subjects from the placebo arm of the COLCORONA study, controlling for sex, age, and 10 principal components for genetic ancestry, with 6,390,776 genetic variants of minor allele frequency ≥ 5%.

**Figure 2.**
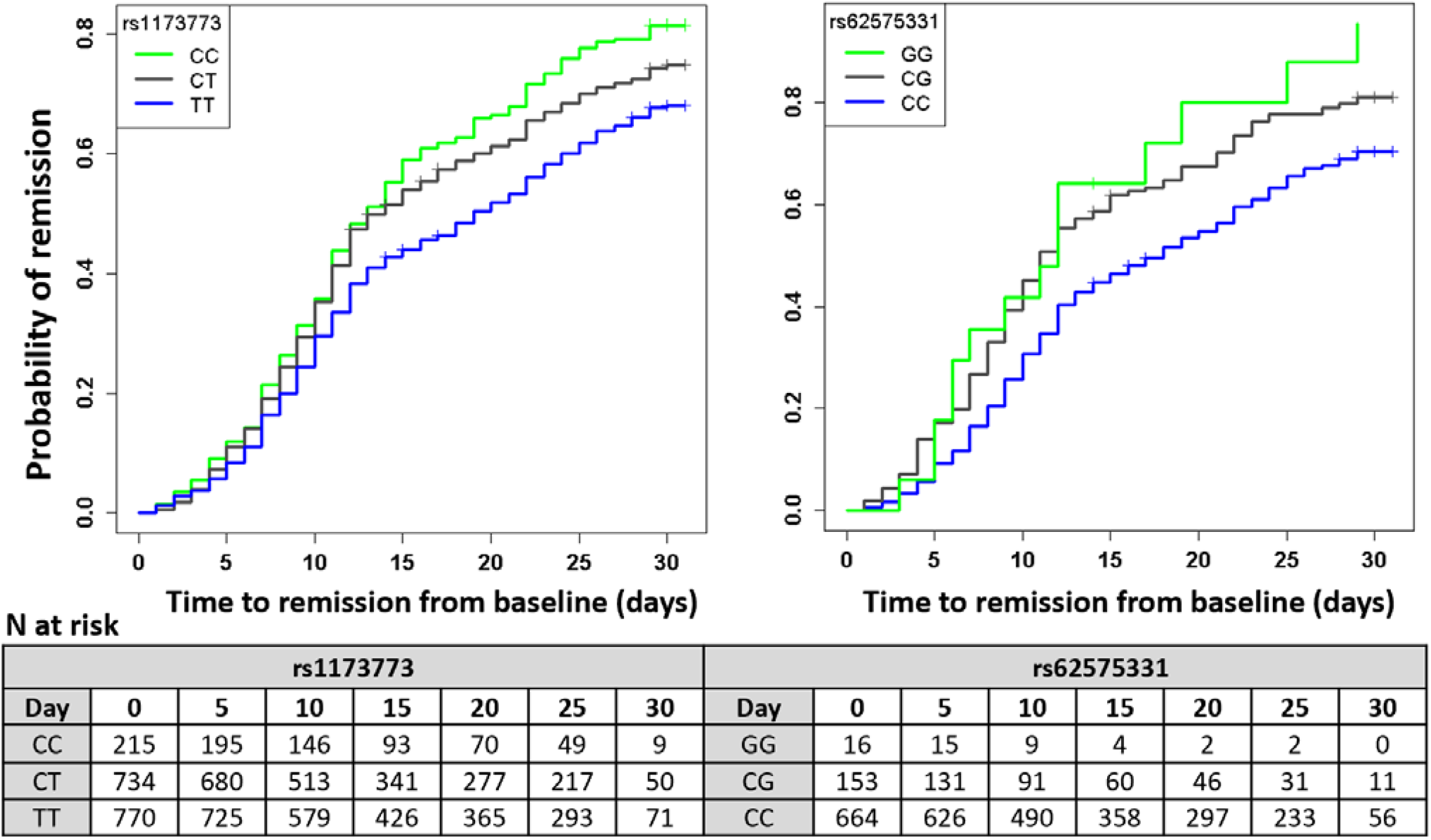
Cumulative incidence curves for COVID-19 symptom remission.

### Results in placebo-treated participants

The most significant association in the GWAS for time to remission of COVID-19 symptoms in the 851 participants from the placebo arm was at 9q33.1 with the intergenic variant rs62575331 (*P =* 2.95 × 10^−8^, MAF = 0.12). When conditioning on rs62575331, no additional genetic variants remained significant at *P <*5 × 10^−8^ in the region and rs62575331 had the highest probability of being causal according to statistical and functional prioritization. The minor allele (G) at the rs62575331 variant leading the 9q33.1 signal was associated with symptom remission (HR=1.57, 95% CI 1.34-1.84). The association of the variant with symptom remission in the colchicine arm was not significant (*P =* 0.15) and the interaction between the variant and colchicine treatment was significant (*P =* 1.19 × 10^−5^), highlighting a possible interaction with colchicine on symptoms duration that may be due to a shared biological pathway between the genetic variant and colchicine (**Table 2**). We calculated that 46%, 62% and 64% of those in the placebo-group with the CC, CG and CG genotypes respectively had symptom remission by day 15 of the study (**Fig. 2, Table 3**). In accelerated failure time modeling, compared to the C allele, the G allele contributed to symptom remission with an acceleration factor of 0.71 (95% CI 0.64-0.80; *P* = 4.83 × 10^−9^). Patients with the rs62575331-GG genotype had symptom remission with an acceleration factor of 0.52 (95% CI 0.36-0.75; *P* = 5.24 × 10^−4^), and patients with the CG genotype an acceleration factor of 0.71 (95% CI 0.61-0.82; *P* = 2.09 × 10^−6^) as compared to the CC genotype. Functional analysis found rs62575331 to be an eQTL of the lncRNA AL355601.1 (GTEx v7; *P* = 1.1 × 10^−18^)^28^, which was found to colocalize with a GWAS of varicose veins.^20,29^ The rs62575331-C allele was associated with height (*P* = 2.3 × 10^−5^), similarly to variants in the pappalysin 1 gene (*PAPPA*) (*P* < 1 × 10^−45^)^22,26,30^ which is the nearest coding gene by transcriptional start site to the leading variant rs62575331. The second-best GWAS signal in the GWAS with the placebo group did not reach statistical significance but was at locus 5p13.3, which was concordant to the region found by GWAS with participants from both study arms. The rs62575331 variant was not associated with the risk of hospitalisation for COVID-19 in the placebo arm (*P =* 0.87) or in the colchicine arm (*P =* 0.61) and had no interaction effect with colchicine on hospitalisation risk.

### GWAS for hospitalisation due to COVID-19

We conducted a GWAS for hospitalisation due to COVID-19 which occurred in 58 (3.1%) of the 1855 subjects included in the analysis. The study had limited power (**Supplementary Methods**) and none of the tested genetic variants passed the GWAS significance threshold (*P <* 5×10^−8^) (**Supplementary Fig. 2**).

## DISCUSSION

We report the results of a genome-wide association study of time to remission of COVID-19 symptoms in an outpatient population recently diagnosed with COVID-19 and who presented with at least one risk factor for COVID-19 complications. We found two genomic regions associated with symptom remission located at 5p13.3 and 9q33.1. The 5p13.3 locus spans the *NPR3* gene, encoding a receptor for the binding of the natriuretic peptides which is involved in the clearance of natriuretic peptides, diuresis, blood pressure, and cardiometabolic diseases.^31^ *NPR3* is involved in the extended renin-angiotensin system which has been proposed as a possible mechanisms involved in the development of lung injury in COVID-19.^32^ The allele associated with remission of COVID-19 symptoms at the chromosome 5 locus has previously been associated with height,^26^ and variants in *NPR3* have also been associated with forced expiratory volume in 1 second (FEV1),^26^ and systolic blood pressure.^27^ The allele associated with symptom remission at the leading variant at 9q33.1 was also previously associated with greater standing height^26^. The variant is an eQTL of a neighbouring lncRNA increasing the risk of venous abnormalities.^20,29^ The chromosome 9 locus variant had a genetic effect on symptom remission which was limited to participants in the placebo arm, and it is possible that the pharmacological effects of colchicine. In the COLCORONA trial, colchicine was not found to modulate duration of COVID-19 symptoms. It is possible that the genetic variant at 9q33.1 may act in a biological pathway that is shared with that of colchicine, which acts on multiple inflammatory pathways including the NLRP3 inflammasome.^33,34^ A recent report by Stella and colleagues^35^ on familial Mediterranean Fever (FMF) in the context of COVID-19 argues that there could be a finely regulated competition between the NLRP3 inflammasome and pyrin that is necessary to maintain protective inflammation levels. FMF is an inherited monogenic autoinflammatory disorder caused by excess activity of the pyrin protein and which is typically treated with colchicine to prevent fever attacks.

The genomic regions identified in this study are novel and do not overlap with previous genetic reports on host genetics of COVID-19 disease. So far, efforts to study the contribution of host genetics to COVID-19 have largely focused on the identification of genetic variants for risk of severe COVID-19 outcomes such as death or hospitalisation for COVID-19 and for genetic variants of risk of infection with the SARS-CoV-2 virus.^3-6^ To the best of our knowledge, ours is the first to study the genetics of COVID-19 symptom remission in an outpatient population. The findings have yet to be replicated using an independent study sample. Concordant with our observations, a survey of the Center for Disease Control has reported that up to 35% of outpatient individuals still had symptoms 14 to 21 days after a COVID-19 diagnosis,^1^ which included young adults without underlying chronic medical conditions. Symptom duration is an important predictor of disease severity which may involve immune responses, as supported by the higher T cell responses in mildly symptomatic individuals with a SARS-CoV2 infection as compared to asymptomatic individuals.^36^ Some patients are experiencing prolonged multiorgan symptoms and complications beyond the initial period of acute infection and illness. A survey led by the UK Government’s Office for National Statistics reported that one in five people who tested positive for COVID-19 had symptoms that lasted for 5 weeks or longer, and one in ten had symptoms that lasted for 12 weeks or longer.^2^ Whether the genetic variants identified in the present study represent risk factors for such long COVID-19 symptoms remains to be verified.

### Limitations

Our study had some limitations. Notably, we had limited power to study the more severe outcomes of death or hospitalisation for COVID-19 due in part to a healthy volunteer bias. The invitation and consent to the genetic substudy occurred after the randomisation visit, and very ill patients may have been less likely to agree to the additional interview or may have already died or been hospitalised by the time of recontact. We made all efforts to rapidly reach all participants and persisted recontact attempts even after the end of the 30-day follow-up period on treatment. Healthy volunteer bias is frequent in optional pharmacogenomic studies of clinical trials. To diminish this bias, a simultaneous consent process with the main study and the collection of genetic material at the randomisation visit is recommended. In practice, however, the additional consent for the genetic study adds time to the recruitment process and can be a deterrent to overall participation, particularly for a disease with acute onset such as COVID-19. Information on symptom types was not collected in this study. Because analyses were conducted with individuals of European genetic ancestry, validation of the genetic associations in other populations will be necessary. Importantly, the results have not yet been replicated in an independent population sample, and we cannot exclude the possibility that the results may be chance findings.

### Conclusion

This is the first study to report a GWAS for time to remission of symptoms in non-hospitalized patients with COVID-19 with at least one risk factor for a severe form of the disease. We found two genomic regions associated with symptom remission located at 5p13.3 and 9q33. In individuals diagnosed with COVID-19. The findings are novel and will need to be replicated in an independent sample.

## Data Availability

The anonymized patient level data from the COLCORONA trial will be shared via the Vivli (vivli.org) data repository. The patient level genetic data underlying this article cannot be shared to preserve the privacy of study participants. Summary statistics from the GWAS results are available for download and visualisation via PheWeb at statgen.org/pheweb/colcorona.

## ACKNOWLEDGMENTS

JCT holds the Canada Research Chair in Personalized Medicine and the Université de Montréal endowed research chair in atherosclerosis. MPD holds the Canada Research Chair in Precision Medicine Data Analysis SdD holds the Université de Montréal Beaulieu-Saucier Chair in Pharmacogenomics. JH is a *Fonds de la Recherche en Santé* (FRQS) Junior 1 fellow. We thank the participants who supported this study. We thank the COLCORONA trial investigators, research nurses, and all research support personnel who made the COLCORONA study possible in a very short period of time. We would like to thank Yannik Couture, Sylvain Versailles, Hugues Gosselin, Marie-Josée Gaulin, Valérie Normand, and Nathalie Zapata for genomic laboratory activities during the pandemic. We are grateful for the research nurses, research personnel and students who contributed to the recruitment and collection of DNA material from study participants: Diane Henry, Rima Amche, Ann Xiuli Chicoine, Hubert Poiffaut, Louis-Éric Lapointe, Isabelle Pinet, Jocelyne Aswad, Mélanie Charron, Frederik Asselin, Audrey Beaulieu, Anne-Marie Gravel, Alexandra Lacroix, Maude Létourneau, Juliette Paul, Laura Provencher, Isabelle Robert, Amélie Desmarais, Sara Hamad, Alexandra Sénécal, Camille Loufti, Simon Olivier Bernard, Martha Lucia Angel, and Meriem Sabir.

## AUTHOR CONTRIBUTIONS

MPD and JCT conceived the experiment; AB, EO, MCG, MCB, MC and MPD conducted statistical analysis; AL, LPLP, GA, SP, JS, MAL and MPD conducted bioinformatics and genetic data analyses; JCT, AD, EG, CS, LMC, NB, RM, ZB, AN, DG, PLLA, GB and DB contributed to data collection; IM, DV, ED, JL, JC conducted laboratory work; MPD wrote the paper with major contributions from JCT, AL, AB, MS, MAL, SdD, and JH. All authors reviewed and approved the manuscript.

## ABBREVIATIONS

CI: Confidence interval
COLCORONA: COLchicine CORONAvirus SARS-CoV-2 trial
COVID-19: Coronavirus disease of 2019
eQTL: expression Quantitative Trait Loci
GWAS: Genome-Wide Association Study
HR: Hazard Ratio
lncRNA: Long non-coding RNA
MAF: Minor allele frequency
OR: Odds ratio

