## Supplemental Material for "Genetics of symptom remission in outpatients with COVID-19"

### SUPPLEMENTARY METHODS

#### *Power calculations*

Power calculations for the GWAS are based on the number of individuals and events observed in the study population and were calculated post-hoc to guide the interpretation of results. There were 1855 participants included in the GWAS for hospitalisation due to COVID-19, of which 58 had an event. The GWAS was analysed using a logistic regression model with a log additive model for genetic variants of minor allele frequency >5% at a $5 \times 10^{-8}$ significance threshold. For genetic variants with minor allele frequencies of 5%, 10%, 15%, 20%, 30% and 40%, we calculated the minimally detectable allelic odds ratio (OR) with 80% power to be 6.5, 4.7, 4.0, 3.7, 3.5, 3.3 respectively using Quanto v1.2.4. There were 1723 participants included in the GWAS for time to remission of COVID-19 symptoms in the placebo and colchicine study arms combined, of which 1254 reached end of symptoms during the 30-day study period and 469 were censored. We used the Cox model power-computing formula based on Hsieh and Lavori method[1] implemented in the SAS POWER procedure (SAS v.9.4). For genetic variants with minor allele frequencies of 10%, 15%, 20%, 30% and 40%, we calculated the minimally detectable allelic hazard ratio (HR) with 80% power to be 1.55, 1.45, 1.40, 1.35, and 1.30 respectively.

#### *Definition of a set of credible risk variants*

We extracted results from the significant regions associated with time to COVID-19 symptoms remission in our GWAS at region 9q33.1, 500kb centered on the top SNP: rs62575331 (chr9:115147521-116147521) and at region 5p13.3 500kb centered on the top SNP: rs1173773 (chr5:32250877-33250877). For the statistical prioritization in the GWAS for time to remission in placebo participants, we considered both 9q33.1 (Locus 1) and 5p13.3 (Locus 2) using the

PAINTOR software. A single locus was used for statistical prioritization in the GWAS for time to remission in both placebo and colchicine arms at 5p13.3 (Locus 3). We defined credible candidate variants (CCVs) as those located within 500kb of the most significant SNP in each locus and with *P* values within two orders of magnitude of this variant. We selected 6, 16 and 31 CCVs for locus 1, locus 2 and locus 3 respectively (**Supplemental Table 2**).

***Conditional analysis***

We used the software GCTA-CoJo[2] to conduct conditional analysis at Locus 1 and Locus 3 (we considered Locus 2 and 3 as equivalent). We performed stepwise model selection on the candidates SNPs and a single independent signal was found in each locus. For Locus 1, rs62575331 was tagged as associated with the outcome with a *P* value from the joint analysis of all the selected SNPs of $5.01 \times 10^{-8}$. For Locus 3, rs1173773 was associated with the outcome with a *P* value from the joint analysis of all the selected SNPs of $6.37 \times 10^{-8}$.

***Statistical prioritization***

We converted GrCh38 coordinates to the previous build referential hg19 and mapped rsID from dbSNP[3] using the R packages *liftover*[4] and *myvariant*[5]. Using PAINTOR (version 3.0)[6], we first performed a multi-locus prioritization with Locus 1 and Locus 2 to identify the most credible causal variants. We used the compiled library of functional annotations provided by PAINTOR's authors to run the PAINTOR model including a large collection of annotations coming from Roadmap/ENCODE data as well as other regulatory and genic annotations. We also did a prioritization using Locus 3 alone. Only the experiment with Locus 3 succeeded to identify a variant with a significant posterior probability of being causal: rs1173773 (pp = 0.86).

Quantification of the enrichment of causal variants within functional classes by PAINTOR estimated the baseline annotation at 3.39, establishing the baseline prior probability for any SNP in the fine-mapping dataset to be causal as 0.033. The most enriched annotation was a chromatin state predicted to have weak transcription in aorta according to data from Bernstein *et al.*[7] using ChromHMM v1.10[8] with a posterior probability of 0.576 and a relative probability to be causal 17.45. The most depleted annotation was a predicted chromatin state of quiescence in H1 BMP4 derived trophoblast cultured cells with a posterior probability of -2.15 and a relative probability to be causal of 0.64. The results are presented in **Supplemental Table 3**.

#### *Functional prioritization*

We used the combination of in silico functional annotations resources from RegulomeDB[9] and DSNetwork[10] for functional prioritization. According to the scoring scheme and the probability of being a regulatory variant[11] estimated by RegulomeDB, the best candidates are the variants rs72764716 (rank = 6, prob = 0.69) and rs1173733 (rank = 5, prob = 1.00) for the Locus 1 and 3 respectively (**Supplemental Table 3**). Regulatory variants were evaluated for their potential deleteriousness using in silico prediction tools provided by DSNetwork. Based on DSNetwork ranking, the top marker is rs62575331 and rs7730564 for Locus 1 (9q33.1) and Locus 3 (5p13.3) respectively (**Supplemental Table 3**).

#### *Selection of the best candidate causal variants*

Based on the statistical and functional prioritization results, we selected the best candidate variants for each of Locus 1 (9q33.1) and Locus 3 (5p13.3). For Locus 1, the highest posterior probability was 0.36 for the top SNP rs62575331. We have considered this analysis as inconclusive. It is possible that the actual causal variants are missing from the set of available variants. The functional prioritization tool enabled to pinpoint rs72764716 (probability of being a regulatory variant of 0.69 in RegulomeDB) and rs62575331 (top SNP in DSNetwork) as credible candidate variants. Those two variants are in perfect LD ($r^2$ = 1, rs72764716(A) allele is correlated with rs62575331(G) allele rs72764716(G) allele is correlated with rs62575331(C) allele and display the same association and effect and we chose the rs62575331 as reference variant for this locus. For Locus 3, the highest posterior probability was 0.86 for the top SNP rs1173773. We have considered this analysis as conclusive. The functional prioritization tool enabled the confirmation of rs1173773 (probability of being a regulatory variant of 1.00 in RegulomeDB) as credible candidate variant and we chose the rs1173773 as reference variant for this locus.

#### *Functional annotation*

We identified and annotated the putative target genes querying the Open Target Genetics Portal[12], GeneMANIA[13], GTEx Portal[14] and PhenoScanner v.2[15,16]. The intergenic variant rs62575331 is associated with the expression of the lncRNA AL355601.1 (ENSG00000234692.1, RP11-445L6.3) in tibial artery tissue. The G allele is associated with an increase of the expression (normalized effect size = 0.59) of this gene ($P = 1.1 \times 10^{-18}$) according to GTEx v7. This lncRNA showed associations with varicose veins and varicose veins of lower extremity.[17] This association is led by the marker rs7469817 whose C-allele is associated with an increase in standing height ($P = 2.6 \times 10^{-14}$), risk of varicose veins ($P = 2.6 \times 10^{-10}$) and forced expiratory volume in 1-second (FEV1) ($P = 2.2 \times 10^{-9}$) among others.[18] Similarly to our candidate variant, this intergenic variant rs7469817 has been found associated with the expression of the lncRNA AL355601.1 (ENSG00000234692.1, RP11-445L6.3) in the tibial artery. The C allele is associated with an increase in the expression (normalized effect size = 0.28) of this gene ($P = 1.1 \times 10^{-18}$) according to GTEx v7 data. Lacking complementary information regarding this non-coding gene, we also included the nearest coding gene *PAPPA* (491,083 bp to canonical TSS) to the set of putative target genes. Variants in *PAPPA* have also been associated with standing height ($P = 1.2 \times 10^{-13}$) and FEV1 ($P = 2.3 \times 10^{-8}$)[18] similarly to the AL355601.1 eQTL rs7469817.

The intronic variant rs1173773 is located in the *NPR3* gene. Using gene expression correlations data from Ensembl,[19] we found that this variant was associated with the expression of several genes with a $P < 0.01$ among which were *MTMR12, SUB1, GOLPH3, TARS* and *NPR3* genes. Most of those genes (4/5) were found associated with anthropomorphic traits such as height and lung functions.[20] In the DIABHYCAR trial (3,126 French Noninsulin-dependent Diabetes, Hypertension, Microalbuminuria or Proteinuria, Cardiovascular Events, and Ramipril), rs1173773 was found to be an independent predictive factor for systolic blood pressure with evidence of modulation by BMI.[21]

Based on functional annotations, we selected two putative target genes: *PAPPA* (Locus 1), *NPR3* (Locus 3) for network analysis with GeneMANIA which highlighted connectivity and support by several common associations in GWAS. Both signals appear to involve similar or closely related phenotypes including height[6,18], systolic blood pressure,[18] and FEV1.[18] Using gene networks between the target genes and genes targeted by the colchicine: (*TUBB, NLRP3* and *NFKB1*), we observed that *PAPPA* was directly connected to the *NFKB1* gene by a co-expression relationship but also through the *C1R* and *IGFBP4* genes. The *PAPPA* gene was also connected to the *NLRP3* gene through the *C1R* gene.

***Colocalization analysis***

To deepen the functional annotation, we performed colocalization analyses between Locus 1 (9q33.1) and Locus 3 (5p13.3) and 1) 16 HGI GWAS results[22], 2) GWAS of time-to-event for 121 outcomes (20 symptoms, 101 CV-related phenotypes) generated by GATE[23,24], 3) GWAS of 36 case-control CV-related phenotypes generated by SAIGE[17] in the context the FinnGen project (Freeze 4), and 4) all cis-eQTL (significant or not) from The Genotype-Tissue Expression (GTEx) Project V7 for relevant tissues: artery (aorta), artery (coronary), artery (tibial), heart (left ventricle) and whole blood.[14] Based, the posterior probabilities of colocalization with Locus 3, we observed eQTLs in the tibial artery for two ncRNA CTD-2203K17.1 and CTD-2066L21.3, and in the aortic artery for CTD-2203K17.1. We also confirmed the putative association with hypertension (H3 = 0.98, in FinnGen and UKB GATE), Hypertensive diseases (H3 = 0.97 in FinnGen). Based, the posterior probabilities of colocalization with Locus 1, we confirmed the putative association with varicose veins (H3 = 0.74 in UKB GATE). No shared signals were found in the HGI studies nor the time-to-event GWAS for different symptoms. The complete results are available in **Supplemental Table 4**.

### SUPPLEMENTARY FIGURES

***Supplementary Figure 1. Flow diagram of participants of the COLCORONA genetic study.***

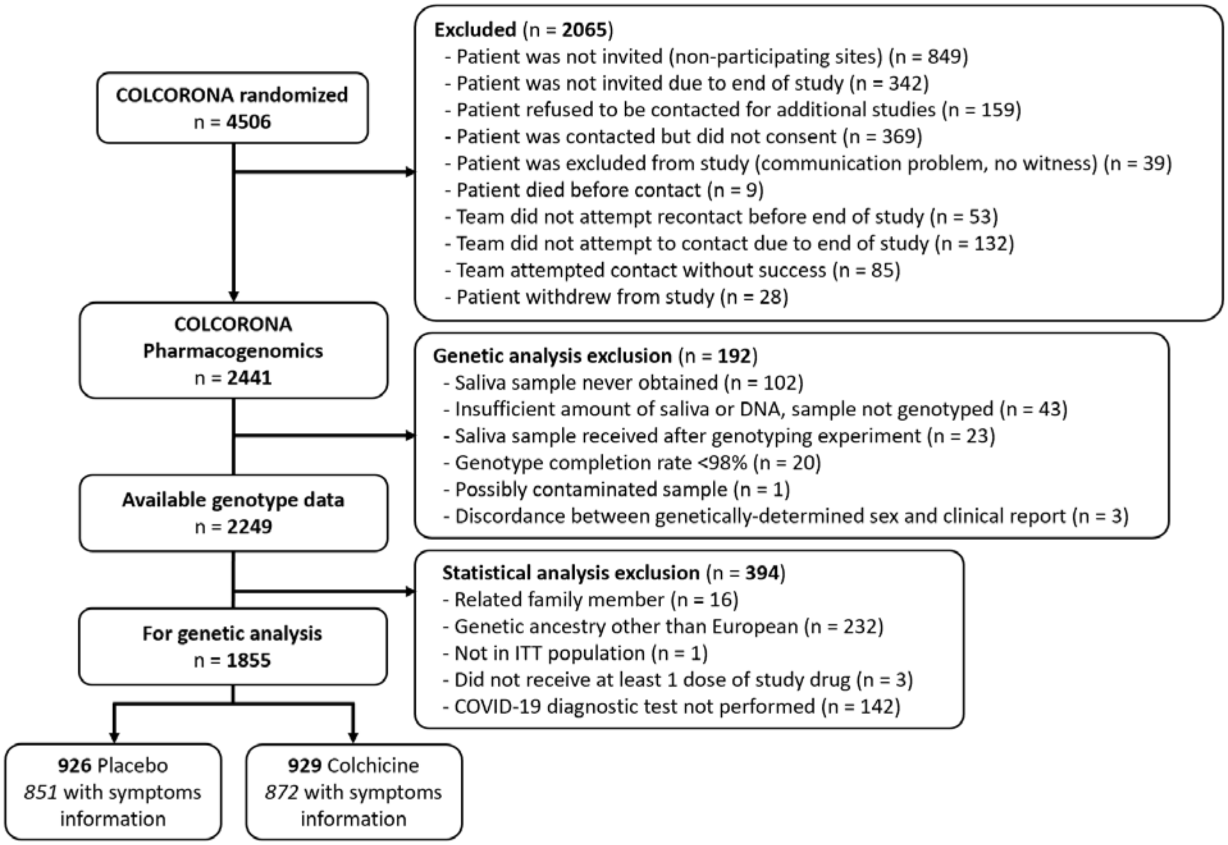

***Supplementary Figure 2. Manhattan plot for the GWAS of hospitalisation for COVID-19***.

Using a logistic regression with 1855 subjects (58 events) from the colchicine and placebo arms of the COLCORONA study, controlling for study arm, sex, age, and 10 principal components for genetic ancestry, with 6,393,360 genetic variants of minor allele frequency ≥ 5%.

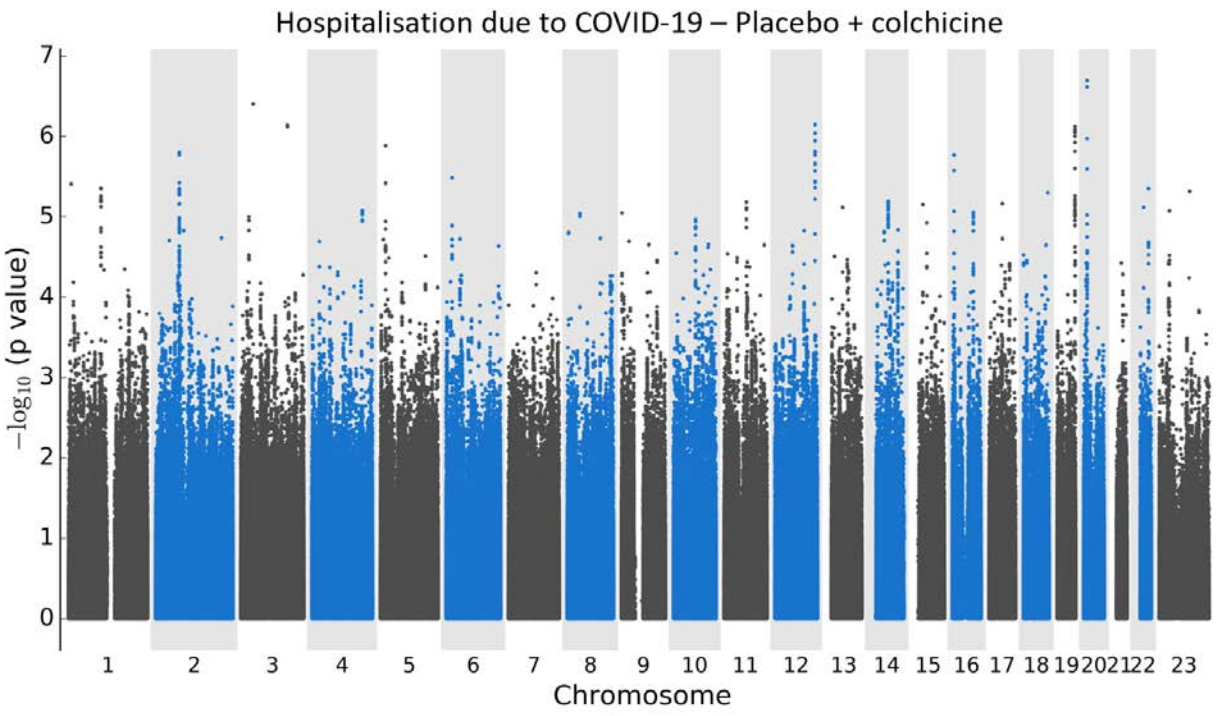

***Supplementary Figure 3. Quantile-quantile (QQ) plots***

a. QQ plot for the GWAS of time to remission of COVID-19 symptoms in 1723 subjects from the colchicine and placebo arms of the COLCORONA study (λ = 1.03).

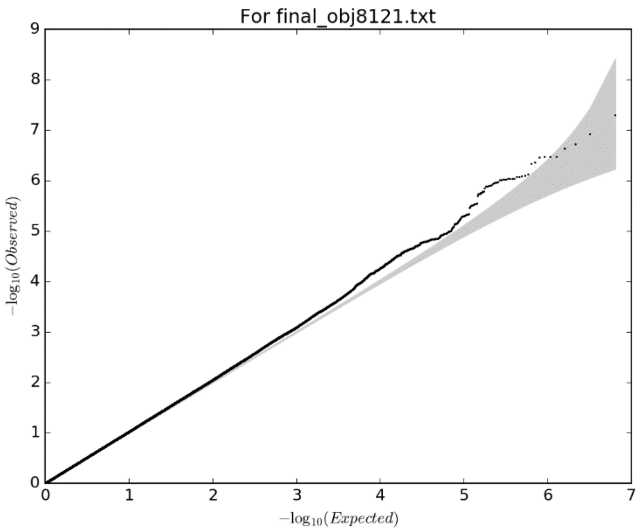

b. QQ plot for the GWAS of time to remission of COVID-19 symptoms in 851 subjects from the placebo arm of the COLCORONA study (λ = 1.04).

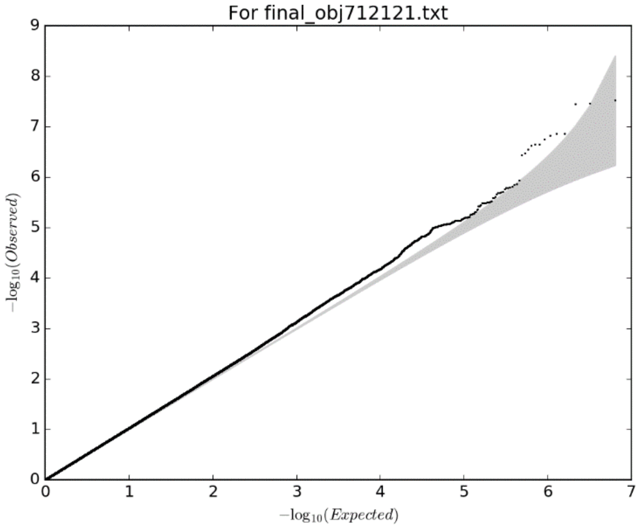

***Supplementary Figure 4. Regional plots for the candidate regions associated with time to remission of COVID-19 symptoms***

**a. The chromosome 5 region.** Plot of association results for genetic variants from the chromosome 5 region (from 32,250,877 to 33,250,877 bp). The degree of linkage disequilibrium ($r^2$) of each genetic variant with rs1173773 estimated from the study population is displayed as blue for [0, 0.2], purple for [0.2, 0.4], green for [0.4, 0.6], orange for [0.6, 0.8], and red for [0.8, 1.0]. Annotations from Ensembl (GRCh38).

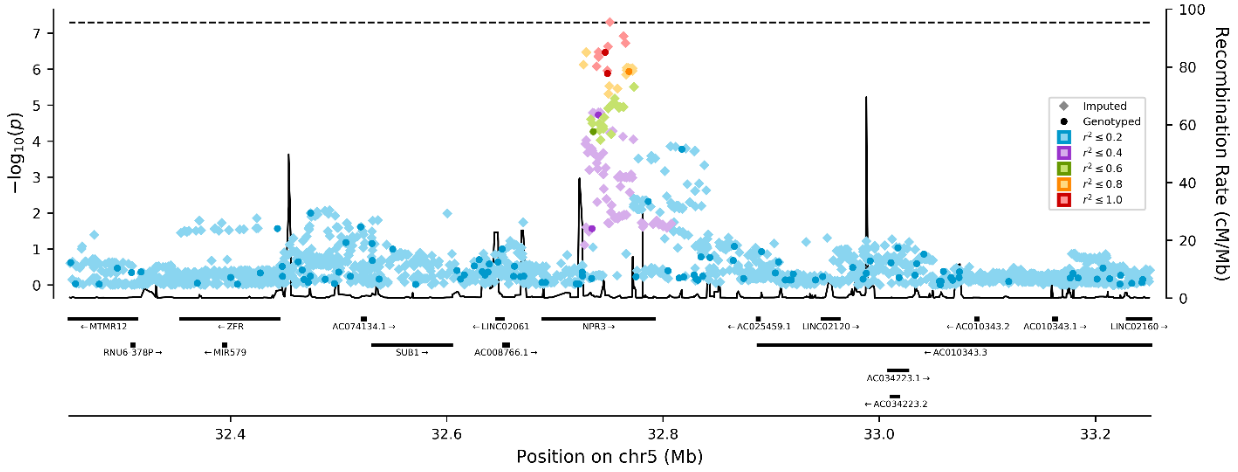

**b. The chromosome 9 region.** Plot of association results for genetic variants from the chromosome 9 region (from 115,147,521 to 116,147,521 bp). The degree of linkage disequilibrium ($r^2$) of each genetic variant with rs62575331 estimated from the study population is displayed as blue for [0, 0.2], purple for [0.2, 0.4], green for [0.4, 0.6], orange for [0.6, 0.8], and red for [0.8, 1.0]. Annotations from Ensembl (GRCh38).

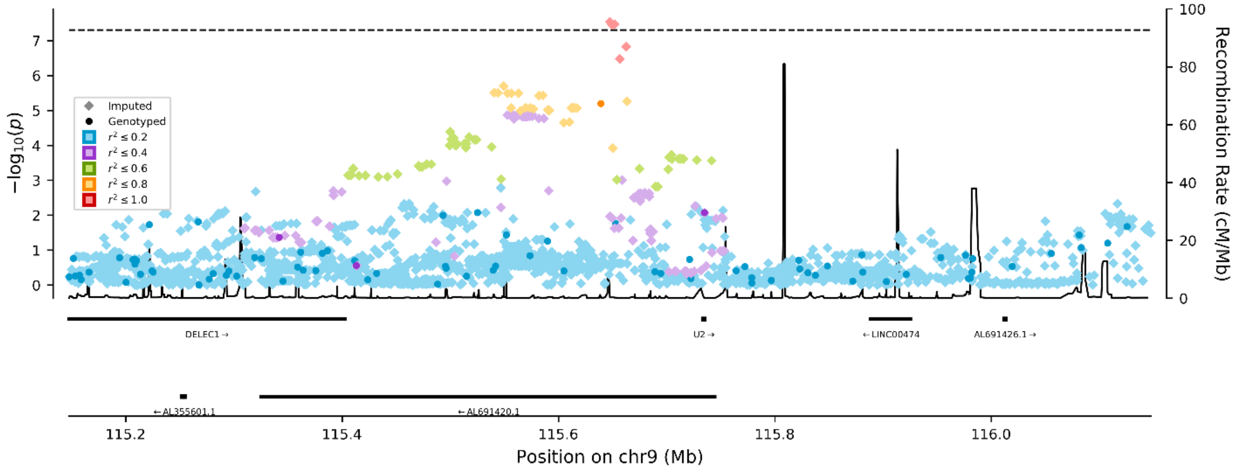

### SUPPLEMENTARY TABLES

#### *Supplementary Table 1. Duration of COVID-19 symptoms by genotype groups*

| | | | Duration of symptoms | | | | | |
|---|---|---|---|---|---|---|---|---|
| | | | All | | Women | | Men | |
| **rs1173773 (chr 5) in placebo + colchicine arms** | | | | | | | | |
| **Genotype** | **Symptoms remission** | **Start date** | **N (%)** | **Mean ± SD** | **N (%)** | **Mean ± SD** | **N (%)** | **Mean ± SD** |
| All | Reported | Baseline | 1252 (72.8%) | 11.8 ± 7.2 | 684 (69.7%) | 12.8 ± 7.2 | 568 (77.0%) | 10.5 ± 6.8 |
| | | Onset | | 17.0 ± 7.9 | | 17.9 ± 8.1 | | 15.9 ± 7.6 |
| | Censored | - | 467 (27.2%) | - | 297 (30.3%) | - | 170 (23.0%) | - |
| TT | Reported | Baseline | 524 (68.1%) | 12.4 ± 7.5 | 284 (66.2%) | 13.5 ± 7.6 | 240 (70.4%) | 11.1 ± 7.2 |
| | | Onset | | 17.9 ± 8.5 | | 19.0 ± 8.8 | | 16.7 ± 7.9 |
| | Censored | - | 246 (31.9%) | - | 145 (33.8%) | - | 101 (29.6%) | - |
| CT | Reported | Baseline | 550 (74.9%) | 11.3 ± 6.9 | 299 (70.5%) | 12.3 ± 6.9 | 251 (81.0%) | 10.1 ± 6.6 |
| | | Onset | | 16.2 ± 7.4 | | 17.0 ± 7.4 | | 15.3 ± 7.5 |
| | Censored | - | 184 (25.1%) | - | 125 (29.5%) | - | 59 (19.0%) | - |
| CC | Reported | Baseline | 178 (82.8%) | 11.5 ± 7.0 | 101 (78.9%) | 12.5 ± 7.2 | 77 (88.5%) | 10.1 ± 6.4 |
| | | Onset | | 16.8 ± 7.4 | | 17.7 ± 7.7 | | 15.6 ± 6.9 |
| | Censored | - | 37 (17.2%) | - | 27 (21.1%) | - | 10 (11.5%) | - |
| **rs62575331 (chr 9) in placebo arm** | | | | | | | | |
| **Genotype** | **Symptoms remission** | **Start date** | **N (%)** | **Mean ± SD** | **N (%)** | **Mean ± SD** | **N (%)** | **Mean ± SD** |
| All | Reported | Baseline | 611 (73.3%) | 11.8 ± 7.2 | 331 (72.0%) | 12.9 ± 7.2 | 280 (75.1%) | 10.4 ± 7.0 |
| | | Onset | | 16.8 ± 7.6 | | 17.8 ± 7.6 | | 15.6 ± 7.5 |
| | Censored | - | 222 (26.7%) | - | 129 (28.0%) | - | 93 (24.9%) | - |
| CC | Reported | Baseline | 471 (70.9%) | 12.2 ± 7.2 | 263 (70.9%) | 13.0 ± 7.1 | 208 (71.0%) | 11.2 ± 7.2 |
| | | Onset | | 17.2 ± 7.7 | | 17.9 ± 7.6 | | 16.4 ± 7.7 |
| | Censored | - | 193 (29.1%) | - | 108 (29.1%) | - | 85 (29.0%) | - |
| CG | Reported | Baseline | 125 (81.7%) | 10.2 ± 6.9 | 61 (75.3%) | 12.4 ± 7.3 | 64 (88.9%) | 8.1 ± 5.9 |
| | | Onset | | 15.4 ± 7.3 | | 17.4 ± 7.5 | | 13.4 ± 6.6 |
| | Censored | - | 28 (18.3%) | - | 20 (24.7%) | - | 8 (11.1%) | - |
| GG | Reported | Baseline | 15 (93.8%) | 10.9 ± 7.6 | 7 (87.5%) | 13.4 ± 9.9 | 8 (100.0%) | 8.6 ± 4.5 |
| | | Onset | | 15.6 ± 7.8 | | 18.1 ± 10.5 | | 13.4 ± 4.0 |
| | Censored | - | 1 (6.3%) | - | 1 (12.5%) | - | 0 (0.0%) | - |
| CG+GG | Reported | Baseline | 140 (82.8%) | 10.3 ± 7.0 | 68 (76.4%) | 12.5 ± 7.5 | 72 (90.0%) | 8.1 ± 5.7 |
| | | Onset | | 15.4 ± 7.3 | | 17.5 ± 7.8 | | 13.4 ± 6.3 |
| | Censored | - | 29 (17.2%) | - | 21 (23.6%) | - | 8 (10.0%) | - |

*Baseline* refers to the COLCORONA study randomisation visit. *Onset* refers to the reported start date of symptoms by the patient; *Reported* refers to patients who reported symptoms remission during the 30-day follow-up period of the study; *Censored* refers to patients who did not report symptoms remission during the 30-day follow-up period of the study. Chr: chromosome; SD: standard deviation.

### SUPPLEMENTARY MATERIAL REFERENCES


1. Hsieh, F.Y. & Lavori, P.W. Sample-size calculations for the Cox proportional hazards regression model with nonbinary covariates. *Control Clin Trials* **21**, 552-60 (2000).
2. Yang, J. *et al.* Conditional and joint multiple-SNP analysis of GWAS summary statistics identifies additional variants influencing complex traits. *Nat Genet* **44**, 369-75, S1-3 (2012).
3. Sherry, S.T. *et al.* dbSNP-database for single nucleotide polymorphisms and other classes of minor genetic variation. *Genome Res* **9**, 677-9 (1999).
4. liftOver: Changing Genomic Coordinate Systems with Rtracklayer. https://bioconductor.org/packages/devel/workflows/vignettes/liftOver/inst/doc/liftov.html .
5. Xin, J. *et al.* High-performance web services for querying gene and variant annotation. *Genome Biol* **17**, 91 (2016).
6. Kichaev, G. *et al.* Improved methods for multi-trait fine mapping of pleiotropic risk loci. *Bioinformatics* **33**, 248-255 (2017).
7. Jones, M.R. *et al.* Ovarian Cancer Risk Variants Are Enriched in Histotype-Specific Enhancers and Disrupt Transcription Factor Binding Sites. *Am J Hum Genet* **107**, 622-635 (2020).
8. Ernst, J. & Kellis, M. Chromatin-state discovery and genome annotation with ChromHMM. *Nat Protoc* **12**, 2478-2492 (2017).
9. Boyle, A.P. *et al.* Annotation of functional variation in personal genomes using RegulomeDB. *Genome Res* **22**, 1790-7 (2012).
10. Lemacon, A. *et al.* DSNetwork: An Integrative Approach to Visualize Predictions of Variants' Deleteriousness. *Front Genet* **10**, 1349 (2019).
11. Dong, S. & Boyle, A.P. Predicting functional variants in enhancer and promoter elements using RegulomeDB. *Hum Mutat* **40**, 1292-1298 (2019).
12. Ghoussaini, M. *et al.* Open Targets Genetics: systematic identification of trait-associated genes using large-scale genetics and functional genomics. *Nucleic Acids Res* **49**, D1311-D1320 (2021).
13. Warde-Farley, D. *et al.* The GeneMANIA prediction server: biological network integration for gene prioritization and predicting gene function. *Nucleic Acids Res* **38**, W214-20 (2010).
14. e, G.P. Enhancing GTEx by bridging the gaps between genotype, gene expression, and disease. *Nat Genet* **49**, 1664-1670 (2017).
15. Staley, J.R. *et al.* PhenoScanner: a database of human genotype-phenotype associations. *Bioinformatics* **32**, 3207-3209 (2016).
16. Kamat, M.A. *et al.* PhenoScanner V2: an expanded tool for searching human genotype-phenotype associations. *Bioinformatics* (2019).
17. Zhou, W. *et al.* Efficiently controlling for case-control imbalance and sample relatedness in large-scale genetic association studies. *Nat Genet* **50**, 1335-1341 (2018).
18. UK Biobank GWAS March 2018 resease http://www.nealelab.is/uk-biobank/.
19. Yates, A.D. *et al.* Ensembl 2020. *Nucleic Acids Res* **48**, D682-D688 (2020).
20. Kichaev, G. *et al.* Leveraging Polygenic Functional Enrichment to Improve GWAS Power. *Am J Hum Genet* **104**, 65-75 (2019).

21. Saulnier, P.J. *et al.* Impact of natriuretic peptide clearance receptor (NPR3) gene variants on blood pressure in type 2 diabetes. *Diabetes Care* **34**, 1199-204 (2011).
22. The COVID-19 Host Genetics Initiative, a global initiative to elucidate the role of host genetic factors in susceptibility and severity of the SARS-CoV-2 virus pandemic. *Eur J Hum Genet* **28**, 715-718 (2020).
23. GATE - UK Biobank Pheweb for browsing the GWAS results of 871 time-to-event phenotypes. http://gate.genohub.org/.
24. Dey, R. *et al.* An efficient and accurate frailty model approach for genome-wide survival association analysis controlling for population structure and relatedness in large-scale biobanks. *bioRxiv*, 2020.10.31.358234 (2020).
